# Clinical Explainability Failure (CEF) & Explainability Failure Ratio (EFR) – changing the way we validate classification algorithms?

**DOI:** 10.1101/2020.08.12.20169607

**Authors:** V. Venugopal, R. Takhar, S. Gupta, A. Saboo, V. Mahajan

## Abstract

Adoption of Artificial Intelligence (AI) algorithms into the clinical realm will depend on their inherent trustworthiness, which is built not only by robust validation studies but is also deeply linked to the explainability and interpretability of the algorithms. Most validation studies for medical imaging AI report performance of algorithms on study-level labels and lay little emphasis on measuring the accuracy of explanations generated by these algorithms in the form of heat maps or bounding boxes, especially in true positive cases. We propose a new metric – Explainability Failure Ratio (EFR) – derived from Clinical Explainability Failure (CEF) to address this gap in AI evaluation. We define an Explainability Failure as a case where the classification generated by an AI algorithm matches with study-level ground truth but the explanation output generated by the algorithm is inadequate to explain the algorithms output. We measured EFR for two algorithms that automatically detect consolidation on chest X-rays to determine the applicability of the metric and observed a lower EFR for the model that had lower sensitivity for identifying consolidation on chest X-rays, implying that trustworthiness of a model should be determined not only by routine statistical metrics but also by novel ‘clinically-oriented’ models.

## Introduction

While there is no doubt that automated diagnosis of pathologies on medical imaging studies using Artificial Intelligence (AI) algorithms will assist physicians in several ways – such as triaging, screening and even reporting of cases – the informed adoption of such algorithms relies inherently on their ‘trustworthiness’. Given the so-called ‘black box’ nature of such algorithms, this trust is built not only by conducting robust validation studies but is also deeply tied to the explainability and interpretability of the algorithm’s output.

Classification algorithms for medical imaging are the most common form of AI algorithms found in the clinical and research domain, and the most common validation metric for reporting the performance of classification algorithms is the Area Under Receiver Operating Curve (AU-ROC), even though its fallacies have been extensively reported(1). Most validation studies on the use of classification algorithms for medical imaging report the performance of the algorithm using image-level or study-level labels, the so-called ground truth (GT)(2). Very few studies on such algorithms report the performance of algorithms on pixel-level or bounding-box labels(3), since such labels are typically reserved for segmentation or lesion detection algorithms.

One potential flaw with studies reporting the performance of classification algorithms is the underlying assumption that in cases where algorithmic output and ground truth match, the reasoning (or explanation) behind the algorithmic output and ground truth are also the same. We test this assumption and define a novel metric – Explainability Failure Ratio.

## Results and Discussion

We define an Explainability Failure as a case where the classification generated by an AI algorithm, for the presence or absence of a pathology, matches with study-level ground truth but the explanation output generated by the algorithm is inadequate to explain the algorithms output.

There are two sequential tests for an Explainability Failure. In the first test, the algorithm fails to localise the pathology correctly and instead localises some other feature on the image. This can be ascertained by comparing the algorithm’s output to bounding boxes or pixel level annotations created by radiologists. Subsequently, we move to the second test, where there is no logical reasoning for failure in the first test that a domain expert can determine. Hence, an Explainability Failure is an instance where even though at study-level a case may be classified as a true positive, there is no discernible explanation for the same. This logical reasoning analysis should include both deductive (finding observations to prove the explanation) & inductive (seeking explanations from the observations) reasoning by a domain expert following the principles of alternative hypothesis. We have documented this approach in our previous work, where we reported the performance of a domain expert’s logical reasoning analysis to understand the outputs of a lung nodule characterization algorithm(4).

The importance of this sequential two-test approach is best highlighted with an example. Consider a validation study of two hypothetical algorithms that can classify brain MRIs into normals and those with infarcts. The ground truth contains pixel level annotations of infarcted areas drawn by radiologists. Heat maps generated by the first algorithm localise the classification output to infarcted areas, whereas maps from the second model localise to an occluded blood vessel. A pure comparison based on localisation of the infarcted area would imply that the second model’s output is an explainability failure, but an expert who tries to reason through the output would infer that the algorithm is in fact localising the occluded blood vessel and hence, is not an explainability failure. On the other hand, had the algorithm not offered a logical reason for its “correctness”, the case would be dubbed an Explainability Failure.

The proposed ability of deep learning algorithms to identify features too subtle or undetectable for the human eyes to see throws up a dilemma – is it right to determine the appropriateness of the explanation of the of an algorithm based on features that are visible only to humans? Examples of such algorithms include those that predict the occurrence of breast cancer and dementia years before they become visible to radiologists on medical images(5,6), and those involved in image reconstruction from raw data. Recognizing this possibility, we call these failures “Clinical” Explainability Failures (CEFs) since the benchmark for explainability of the algorithms is rooted in clinical knowledge and science. The ratio of the number of CEFs to the total number of true positives in a test dataset can give an indication of the prevalence of such failures and can hence be dubbed the Explainability Failure Ratio (EFR).

In a recent study that evaluated the trustworthiness of saliency maps for localizing abnormalities in X-rays, the authors have used localization and explainability interchangeably(7). We believe it is important to make a distinction between explainability & localization especially if the classification or characterisation of a detected object (or lesion, in medical imaging) is not possible without the knowledge of its environment. A dense lesion in the lungs can sometimes be difficult for a radiologist to differentiate from collapse (atelectasis) & consolidation by just localizing the lesion itself. Their interpretation may depend on ancillary findings like tracheal shift and volume loss, which help drive the decision towards a specific class. Similarly, there is a possibility that neural networks are identifying patterns correlated with a pathology class that may not be inherently apparent to human readers. The saliency maps generated might be highlighting patterns and associations learnt from the training data which might not localize to the lesion itself.

## Methods – Evaluation

### Models

We considered two different chest X-ray classification models for this evaluation – a pneumonia detection algorithm developed by Cadrin-Chênevert et al(8) and an open-sourced reimplementation of Stanford’s baseline X-Ray classification model (Modified_CheXNeXt or M.CheXNeXt).

### Pneumonia Detection & Classification

The pneumonia detection algorithm consists of 2 pipelines to detect and classify lung opacities on any frontal chest X-ray. The detection pipeline consists of an ensemble of 5 models trained using R-FCN, Relation Network, and RetinaNet on the various resolutions of images. It outputs bounding boxes on located lung opacities with confidence greater than 0.3 by default. The classification pipeline classifies a frontal chest X-ray into pneumonia and non-pneumonia.

### M.CheXNeXt

The M.CheXNeXt model is an open-sourced re-implementation of Stanford’s baseline X-Ray classification model which uses DenseNet121 as its backbone architecture. It classifies any X-ray (Frontal or Lateral) into 14 different classes. The model was trained with the same configuration as described in the CheXNeXt article(9). It also provides heat maps corresponding to each class to indicate the confidence of the detected disease using guided GRAD-CAM(10).

## Evaluation

### Dataset

To evaluate the applicability of the metric, a comparative evaluation of the two models described above was performed on a dataset containing 611 frontal chest X-ray studies with study-level Ground-Truth (GT) created by a radiologist with 10 years’ experience of thoracic radiology. The dataset contained 189 (30.9%) normal X-rays, 157 (25.7%) with consolidation and 265 (43.4%) with other pathologies. Images with consolidation had additional bounding box GT to localise the pathology on each such image.

### Clinical Explainability Failure Evaluation

The outputs generated by the two models on the dataset above were first compared to the image-level GT for consolidation at the model output threshold with highest Mathew’s Correlation Coefficient(11). Subsequently, true positive cases were subjected to the two tests for explainability failures described before, by comparing the localisation output generated by the models to the bounding box GT by the radiologist. To compare the outputs of the two models, the localisation outputs generated by the M.CheXNeXt model were converted to bounding boxes in a way such that the bounding box covers the heat map completely.

### Estimation of Localisation Accuracy

Two common metrics for estimating the accuracy of localisation algorithms are DICE Score and Jaccard Index (7). When these metrics are reported in aggregate, they tend to normalise the failures of individual instances – a significant drawback when clinically validating a localisation algorithm. An additional problem is faced when there is a mismatch between the number of ‘locations’ produced by an algorithm and pixel-level expert annotations. Hence, we applied a different approach by creating a greedy matching algorithm that picks each bounding box GT annotation for each true positive case and matches it to the nearest overlapping model generated bounding box on the X-ray image. A match is considered positive when the bounding box GT overlaps with one of the top three (as per their probability scores) model-generated bounding box. If one or many model-generated bounding boxes are completely enclosed within a single GT bounding box, or vice-versa, a perfect match is considered. Once this matching process is completed, cases with unmatched GT bounding boxes were evaluated by a radiologist for logical reasoning analysis. Upon failing in the logical reasoning process, the case was labelled as a CEF and the model’s EFR calculated.

### Results

The Pneumonia Detection & Classification algorithm had a sensitivity of 57% at a threshold of 0.28 correctly classifying 90 of the 157 cases with consolidation. Of these 90 cases only 2 cases of CEF were observed giving an EFR of 2.2%. M.CheXNeXt had a sensitivity of 76% at a threshold of 0.06 correctly classifying 120 of the 157 cases with consolidation. Of these 120 cases, 16 CEFs were observed yielding an EFR of 13.3%. **Figure 1** shows the complete workflow for performing a CEF analysis, along with examples of CEFs from our study. All the images, along with their model generated and radiologist-generated bounding boxes are available at this link.

**Figure 1.**
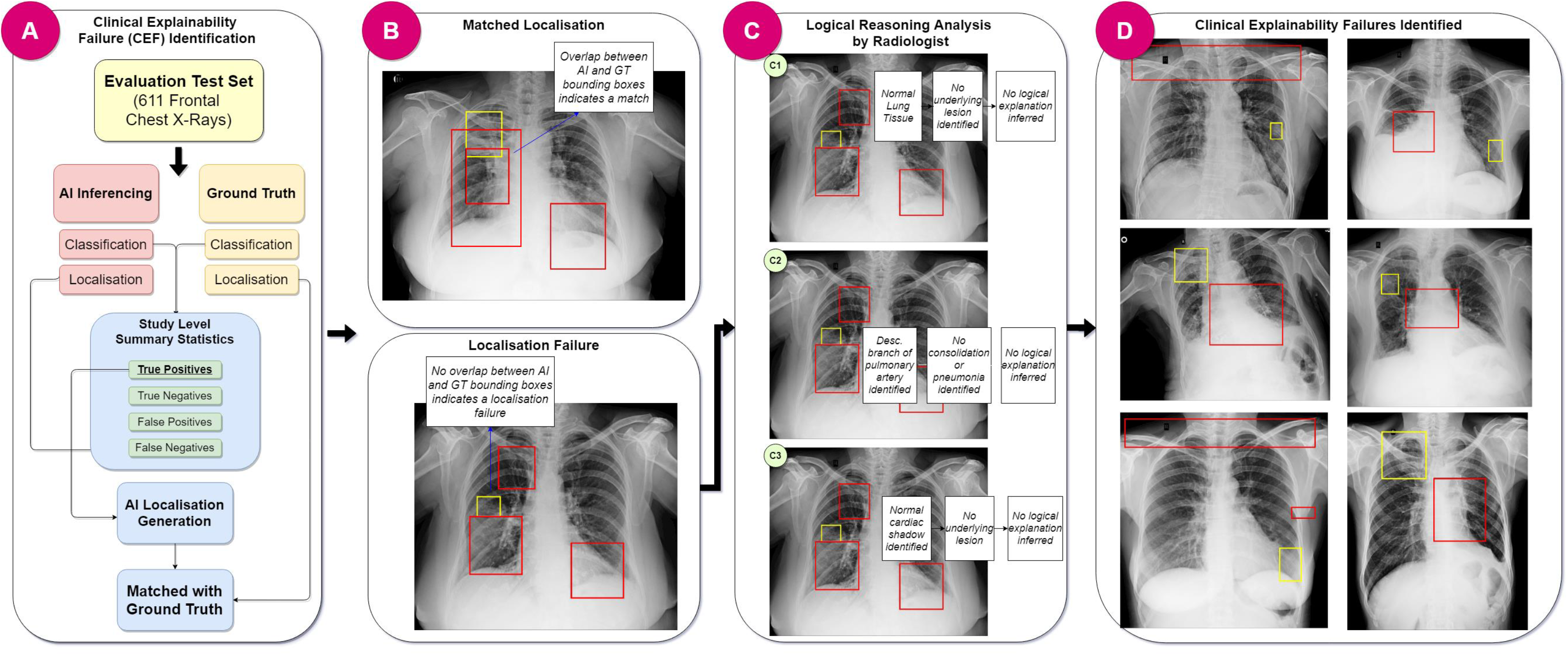
Clinical Explainability Failure Analysis. illustrates the process of Clinical Explainability Failure (CEF) analysis by a radiologist. In the above images, red rectangles denote AI generated localisation outputs in the form of bounding boxes and yellow rectangles denote radiologist annotated bounding boxes (Ground Truth). **Section A** shows the study pre-requisites and subsequent summary statistics calculation. Note that AI localisations are generated for True Positives and compared to Ground Truth localisations for the same. **Section B** shows localisation matching process. **Section C** shows the logical reasoning analysis of a single case wherein the radiologist tries to infer rationale for each of the AI generated bounding boxes. **Section D** shows examples of Clinical Explainability Failures from the CheXNeXt model - each image was classified as having ‘Consolidation’ by the model but failed in both the localisation and logical reasoning analysis.

From these results we can infer that even though the Pneumonia Detection & Classification algorithm had a lower sensitivity than M.CheXNeXt, its Explainability Failure Ratio was significantly lower, and can hence postulate that the Pneumonia Detection algorithm is the more explainable of the two.

## Conclusion

Standard statistical metrics do not capture this ‘clinical’ nuance, missing out on which can have potentially dangerous consequences. It is critical to not only know ‘why’ a machine learning or deep learning model is giving a particular output, but also to concretely determine whether it was logical in its ‘reasoning’. It would be prudent for all studies reporting the performance of a classification algorithm to report such failures – such failures can provide important insights into the trustworthiness of an algorithm especially since, by definition, these cases are a subset of true positives and, in our experience, true positives are the most ignored part of a validation study. While our previous work on algorithmic auditing recommended diving deep into false positive and false negative cases(12), we now additionally recommend diving deep into true positives. True positive cases are generally celebrated as a success of such models – we demonstrate that in a significant number of such cases, the rationale behind the model’s output is unfathomable to human experts, and apparently incorrect.

## Data Availability

Chest X-Ray data with bounding boxes drawn by AI and humans is available at https://github.com/caringresearch/clinical-explainability-failure-paper/

## Acknowledgments

We would like to thank members of the CovBase (covbase.igib.res.in) initiative for helping spark discussion about this concept during its initial stages. We also thank Dr. Alexandre Cadrin-Chenevert for providing his Pneumonia Detection & Classification algorithm which was the winner of the RSNA-2018 Kaggle Chest X-Ray challenge.

